# Association of alpha-aminoadipic acid (2-AAA) with cardiometabolic risk factors in healthy and high-risk individuals

**DOI:** 10.1101/2023.06.05.23290990

**Authors:** Stacy Desine, Curtis L. Gabriel, Holly M. Smith, Olivia R. Antonetti, Chuan Wang, M. Wade Calcutt, Amanda C. Doran, Heidi J. Silver, Sangeeta Nair, James G. Terry, J. Jeffrey Carr, MacRae F. Linton, Jonathan D. Brown, John R. Koethe, Jane F. Ferguson

**Affiliations:** Division of Cardiovascular Medicine, Vanderbilt University Medical Center, Nashville, TN, USA; Division of Gastroenterology, Hepatology and Nutrition, Vanderbilt University Medical Center, Nashville, TN, USA; Tennessee Center for AIDS Research, Vanderbilt University Medical Center, Nashville, TN, USA; Department of Biochemistry, Mass Spectrometry Research Center, Vanderbilt University, Nashville, TN, USA; Department of Radiology and Radiological Sciences, Vanderbilt University Medical Center, Nashville, TN, USA; Division of Infectious Diseases, Vanderbilt University Medical Center, Nashville, TN, USA

## Abstract

Plasma levels of the metabolite alpha-aminoadipic acid (2-AAA) have been associated with risk of type 2 diabetes (T2D) and atherosclerosis. However, little is known about the relationship of 2-AAA to other cardiometabolic risk markers in pre-disease states, or in the setting of comorbid disease. We measured circulating 2-AAA using two methods in 1) a sample of 261 healthy individuals (2-AAA Study), and 2) in a sample of 134 persons comprising 110 individuals with treated HIV, with or without T2D, a population at high risk of metabolic disease and cardiovascular events despite suppression of circulating virus, and 24 individuals with T2D without HIV (HATIM Study). We examined associations between plasma 2-AAA and markers of cardiometabolic health within each cohort. We observed differences in 2-AAA by sex and race in both cohorts, with higher levels observed in men compared with women, and in Asian compared with Black or white individuals (P<0.05). There was no significant difference in 2-AAA by HIV status within individuals with T2D in the HATIM Study. We confirmed associations between 2-AAA and dyslipidemia in both cohorts where high 2-AAA associated with low HDL cholesterol (P<0.001) and high triglycerides (P<0.05). As expected, within the cohort of people with HIV, 2-AAA was higher in the setting of T2D compared to pre-diabetes or normoglycemia (P<0.001). 2-AAA was positively associated with body mass index (BMI) in the 2-AAA Study, and with waist circumference and measures of visceral fat volume in HATIM (all P<0.05). Further, 2-AAA associated with increased liver fat in persons with HIV (P<0.001). Our study confirms 2-AAA as a marker of cardiometabolic risk in both healthy individuals and those at high cardiometabolic risk, reveals relationships with adiposity and hepatic steatosis, and highlights important differences by sex and race. Further studies are warranted to establish molecular mechanisms linking 2-AAA to disease in other high-risk populations.

## INTRODUCTION

Cardiometabolic diseases, including diabetes and cardiovascular disease (CVD) are increasing in prevalence globally and represent a major contributor to mortality (Tsao et al., 2022). Known risk factors include obesity, dyslipidemia, dysregulated glucose metabolism and inflammation (Shah et al., 2018). However, after accounting for these risk factors there remains a high degree of variability in disease susceptibility, and a clear need for more refined biomarkers of cardiometabolic risk to improve our understanding of the underlying disease mechanisms and to improve prediction and treatment of at-risk individuals.

Cardiometabolic diseases are characterized by changes in metabolism that may contribute to disease pathophysiology, or may act as biomarkers of disease progression (Upadhyay, 2015). Circulating metabolites that associate with disease states can shed light on underlying disease etiology, biological mechanisms, and may have clinical utility for prediction (Chu et al., 2021). Strategies to identify individuals at high cardiometabolic risk and to modulate disease processes in these individuals before onset of overt disease, would have significant impact in reducing mortality, morbidity, and healthcare costs. For this approach to be successful, early biomarkers of disease that predict at-risk individuals are required, as well as discovering novel pathways for therapeutic targeting. To this end, studying both healthy individuals, as well as individuals with conditions that place them at higher risk of cardiometabolic diseases, may provide an important model to identify novel physiologic relationships.

The metabolite alpha-aminoadipic acid (2-AAA) is associated with the development of type 2 diabetes (T2D) (Wang et al., 2013) and atherosclerosis (Saremi et al., 2017), potentially identifying at-risk individuals before development of other known risk markers (Lee et al., 2019). Relatively little is known about the function of 2-AAA, or potential mechanisms linking 2-AAA to disease. 2-AAA is derived from the breakdown of the essential amino acid lysine, and is primarily metabolized within mitochondria, with potential involvement in oxidative stress (Estaras et al., 2020; Luna et al., 2021). Elevated 2-AAA is associated with increased insulin secretion, obesity, and dysregulated mitochondrial metabolism (Wang et al., 2013, 2021; Wu et al., 2014; Ho et al., 2016; Plubell et al., 2018; Lee et al., 2019). This makes 2-AAA an interesting novel candidate in cardiometabolic disease biology. However, the relationships between 2-AAA and other cardiometabolic risk markers have not been well-described.

The purpose of this study was to characterize the association between 2-AAA and other demographic and circulating markers in a sample of healthy individuals, as well individuals at high risk of metabolic and cardiovascular disease. As chronic viral infections, including treated human immunodeficiency virus (HIV), predispose individuals to a higher incidence of cardiometabolic disease and earlier onset, these conditions can serve as an models of exaggerated or accelerated risk to further identify important physiologic relationships (Barale et al., 2022; Gooden et al., 2022; Rivera et al., 2022; Spieler et al., 2022). Here, we assess the relationship of 2-AAA with range of cardiometabolic disease conditions and risk factors among healthy individuals and those with treated HIV infection.

## MATERIALS AND METHODS

### Study Populations

Samples and data from two independent studies are included here. Participants of both studies were recruited from the same geographic area (Nashville, TN, and surrounding areas), and study procedures completed at Vanderbilt University Medical Center.

#### Determinants of 2-AAA: Screening Study (2-AAA Study)

Healthy adults (non-pregnant and non-lactating women and men, age 18-45 years) were recruited to complete a single study visit as part of a cross-sectional study at Vanderbilt University Medical Center between November 2018 and June 2021. Exclusion criteria included body mass index (BMI) >30 kg/m^2^, active use of tobacco products, active use of prescription medications (apart from hormonal birth control), and diagnosis of diabetes mellitus, cardiovascular disease, renal disease, liver disease, or bleeding disorders. Data for 261 individuals who completed study procedures (vital signs, anthropometric measurements), provided a fasting blood sample, and had sufficient plasma available for 2-AAA measurement are included in the current analysis. All participants provided written, informed consent, and the study was approved by the Vanderbilt University Institutional Review Board.

#### The HIV, Adipose Tissue Immunology, and Metabolism Study (HATIM) Study

Adults with human immunodeficiency virus (HIV, N=112) were recruited from the Vanderbilt Comprehensive Care Clinic between August 2017 and November 2019. Participants were on combination antiretroviral therapy (ART) for ≥18 months, with a minimum of 12 months of sustained suppression of plasma viremia at enrollment and had no known inflammatory or rheumatologic conditions. Exclusion criteria were self-reported heavy alcohol use (>11 drinks/week), known cirrhosis, active hepatitis B or C, cocaine or amphetamine use, and use of corticosteroids or growth hormones. By design and to enrich for the presence of cardiometabolic disease, the cohort enrolled approximately equal numbers of individuals who were normoglycemic (HbA1c < 5.7 or fasting blood glucose (FBG) < 100 mg/dL); pre-diabetes (HbA1c 5.7%-6.4% and/or FBG 100-126 mg/dL); and diabetes (HbA1c ≥ 6.4%, and/or FBG ≥ 126 mg/dL or on diabetes medication). To allow for direct comparison of 2-AAA levels with HIV-negative individuals, the study also recruited individuals with diabetes but without HIV (N=24). Participants provided written, informed consent, and the study was approved by the Vanderbilt University Institutional Review Board (ClinicalTrials.gov Identifier: NCT04451980).

### Measurement of 2-AAA

In the *2-AAA Study*, plasma levels of 2-AAA were quantified by liquid chromatography mass spectrometry (LCMS) at the Vanderbilt Mass Spectrometry Core. Samples were spiked with internal standard (Arginine-15N4, Sigma Aldrich), extracted with methanol, and derivatized with dansyl chloride (Sigma Aldrich) prior to analysis. The dansyl derivative of 2-AAA ([M+H]+ 395.1271) was measured by targeted selected ion monitoring (SIM) using a Vanquish ultrahigh performance liquid chromatography (UHPLC) system interfaced to a QExactive HF quadrupole/orbitrap mass spectrometer (Thermo Fisher Scientific). Data acquisition and quantitative spectral analysis were conducted using Thermo-Finnigan Xcaliber version 4.1 and Thermo-Finnigan LCQuan version 2.7, respectively. Calibration curves were constructed by plotting peak area ratios (2-AAA / Arg-15N4) against analyte concentrations for a series of 2-AAA standards. Electrospray ionization source parameters were tuned and optimized using an authentic 2-AAA reference standard (Sigma Aldrich) derivatized with dansyl chloride and desalted by solid phase extraction prior to direct liquid infusion.

In the *HATIM Study*, plasma 2-AAA was measured as part of a metabolomics panel, at the Southeast Center for Integrated Metabolomics (SECIM) at the University of Florida, using previously described methods (O’Kell et al., 2017, 2019). Briefly, plasma samples were spiked with internal standards solution. Proteins were precipitated using 8:1:1 Acetonitrile: Methanol: Acetone (Fisher Scientific, San Jose, CA), and the supernatant dried under a gentle stream of nitrogen at 30°C (Organomation Associates, Inc., Berlin, MA). Samples were reconstituted with injection standards solution. LC-MS untargeted metabolomics was performed on a Thermo Q-Exactive Orbitrap mass spectrometer equipped with a Dionex UPLC system (Thermo, San Jose, CA). Percent relative standard deviation of internal standard peak areas were calculated to evaluate extraction and injection reproducibility. Mzmine 2 was used to identify features, deisotope, align features and perform gap filling. The data was searched against SECIM internal retention time metabolite library. All adducts and complexes were identified and removed from the data set. Ion counts from features mapping to alpha-aminoadipic acid in positive ion mode were summed for analysis. Because measurement of 2-AAA was conducted at different sites, studies were analyzed separately.

### Lipid and Biomarker Measurement

In the *2-AAA Study*, serum lipids were profiled at the Vanderbilt Lipid Laboratory. Briefly, total cholesterol and triglycerides (TG) were measured by standard enzymatic assays. High-density lipoprotein (HDL) was measured with the enzymatic method after precipitation of VLDL and LDL using polyethylene glycol reagent (PEG). LDL cholesterol was calculated using the Friedewald equation (Friedewald et al., 1972). In the *HATIM Study*, fasting plasma HDL, LDL, and TG were measured using the selective enzyme hydrolysis method (Abbott, Chicago, IL). In the 2-AAA Study, fasting glucose was measured at the study visit by finger prick (AimStrip Plus Blood Glucose Meter, Germaine Laboratories Inc., San Antonio TX). In the HATIM Study, insulin was measured by radioimmunoassay (Millipore Cat. # PI-13K). The assay utilizes ^125^I - labeled insulin and a double antibody/PEG technique to determine serum insulin levels. The assay was modified by the Vanderbilt Hormone and Analytical Services Core to improve the sensitivity to 1uU/ml(0.04ng/ml). Glucose and hemoglobin A1c (HbA1c) were measured in fasting blood samples at the Vanderbilt Clinical Chemistry Laboratory.

### Body Composition Analysis

In the *HATIM Study,* individuals underwent computed tomography (CT) imaging using a Siemens Somatom Force multidetector scanner (Erlangen, Germany) to acquire chest, abdominal and liver images, as described (Gabriel et al., 2021; Bailin et al., 2022). Briefly, separate non-contrast electrocardiogram-gated thorax (top of the aortic arch through the lung base) and abdominal (diaphragm to lumbosacral junction) scans were performed using a scanning protocol and image interpretation approach previously described (Carr et al., 2005; VanWagner et al., 2014; Terry et al., 2017). Abdominal subcutaneous adipose tissue (SAT) and visceral adipose tissue (VAT) volumes were measured within a 10-mm block of images consisting of eight images, 1.25-mm thick, at the L4-5 vertebrae using Osirix software. Pericardial adipose tissue (PAT) volume was measured within a 45-mm block of images spanning 15 mm above and 30 mm below the superior extent of the left main coronary artery, which includes the adipose tissue located around the epicardial coronary arteries (left main coronary, left anterior descending, right coronary, and circumflex arteries) as well as the epicardial and PAT around the coronary arteries (Alman et al., 2016; Miljkovic et al., 2020). Images at T12-L1 were used to identify the liver below the right diaphragm corresponding to superior aspects of the right and medial lobes or hepatic segments 4a, 7, and 8 using the Couinaud classification system. Three regions of interest within homogenous portions of the liver at three levels were identified and liver density was averaged from the nine total regions. Tissue radiodensity was quantified using the Hounsfield Units scale where water has a value of 0 HU and air has a value of −1000 HU.

### Statistical Analysis

Plasma 2-AAA was assessed for normality of distribution through visualization, and testing for skewness and kurtosis, and was found to follow a normal distribution in both the 2-AAA and HATIM studies. Two individuals were considered outliers for 2-AAA in HATIM (>3 SD from the mean) and were removed prior to analysis. Associations between 2-AAA and continuous variables were analyzed using linear regression models. Analyses between 2-AAA and discrete variables were analyzed by T-test or ANOVA. Models were adjusted for sex and race in both studies and for additional covariates in HATIM (smoking, diabetes group). Models were further adjusted for other risk factors as indicated in the corresponding results sections, including BMI, cholesterol, HDL, LDL, TG, fasting glucose. P<0.05 was considered statistically significant, and Bonferroni P<0.05 considered statistically significant for post hoc multiple testing correction. Analyses were completed and results visualized using IBM SPSS Statistics version 28 (IBM, Armonk NY) and GraphPad Prism version 9.4.1 (GraphPad Software, San Diego, CA).

## RESULTS

The characteristics of the participants of the 2-AAA Study are shown in **Table 1**. Characteristics of the participants of the HATIM Study are shown in **Table 2**. Participants of the 2-AAA study were 72% female, and 74% white, with an average age of 28 years. Participants of the HATIM study were 67% male, and 54% white, with an average age of 48 years. Plasma 2-AAA in persons with HIV (PWH) with diabetes (ion count 312×10^4^±75×10^4^) was slightly higher than that in HIV-negative with diabetes (ion count 271×10^4^±74×10^4^), but the difference was not statistically significant (P=0.08).

**Table 1.** Characteristics of the participants of the 2-AAA Screening Study

|  | **Male (N=72)** | **Female (N=189)** |
| --- | --- | --- |
|  | **Mean (SD)** | **Mean (SD)** |
| **Age (years)** | 28.96 (6.92) | 27.76 (7.2) |
| **Race (N Black, white, Asian, other)** | 3, 55, 8, 6 | 14, 139, 25, 11 |
| **BMI (kg/m<sup>2</sup>)** | 24.77 (2.9) | 22.94 (2.9) |
| **Systolic Blood Pressure (mmHg)** | 120.57 (13.5) | 111.26 (10.7) |
| **Diastolic Blood Pressure (mmHg)** | 73.80 (9.5) | 69.67 (8.1) |
| **Glucose (mg/dL)** | 90.0 (7.6) | 91.18 (8.2) |
| **Total cholesterol (mg/dL)** | 166.56 (30.7) | 167.94 (32.4) |
| **HDL (mg/dL)** | 53.47 (10.7) | 64.67 (13.1) |
| **LDL (mg/dL)** | 95.56 (24.9) | 87.41 (24.5) |
| **TG (mg/dL)** | 87.44 (37.5) | 79.29 (37.1) |
| **2-AAA (ng/ml)** | 95.99 (33.8) | 68.44 (27.7) |

**Table 2.**
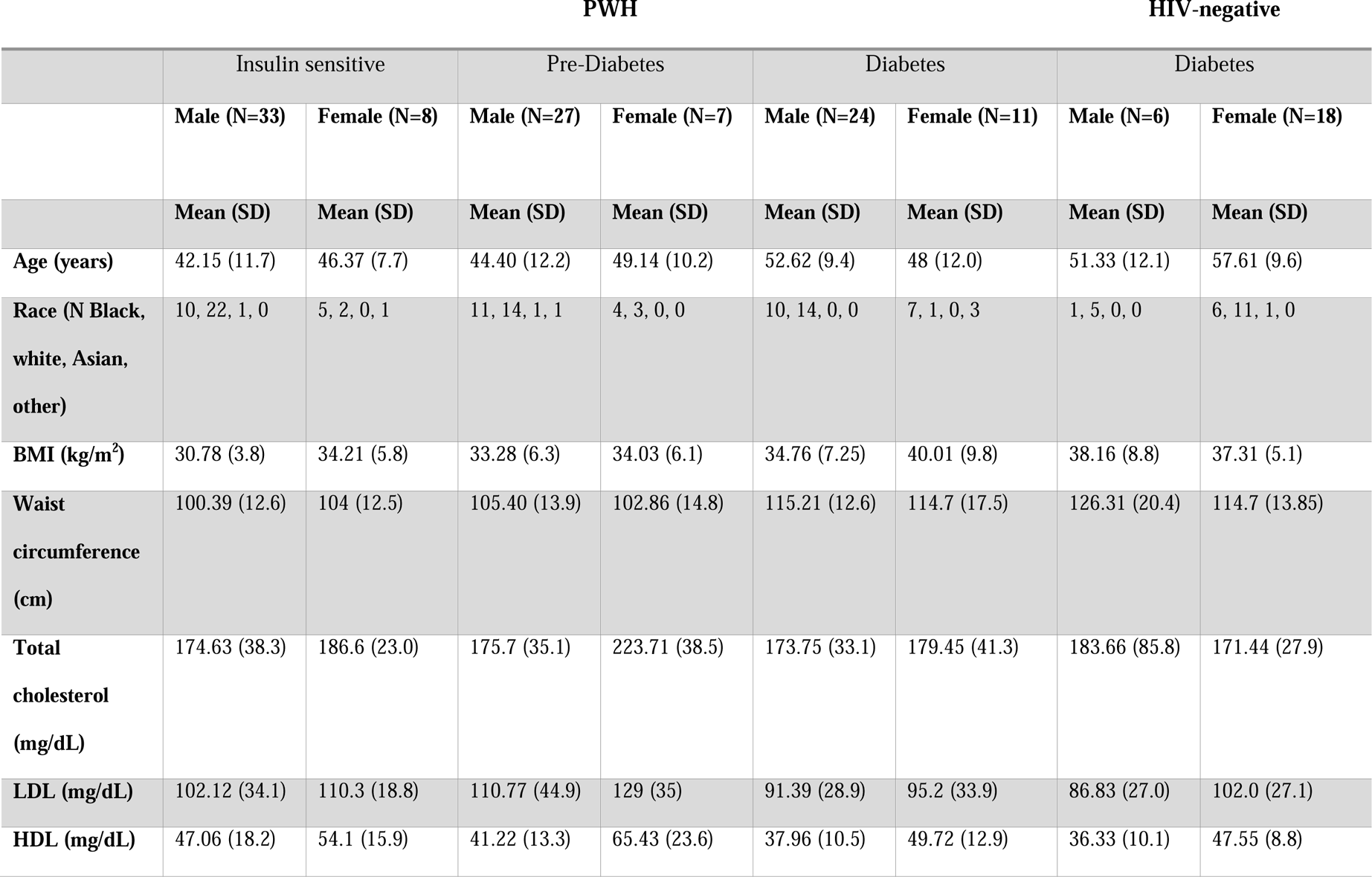
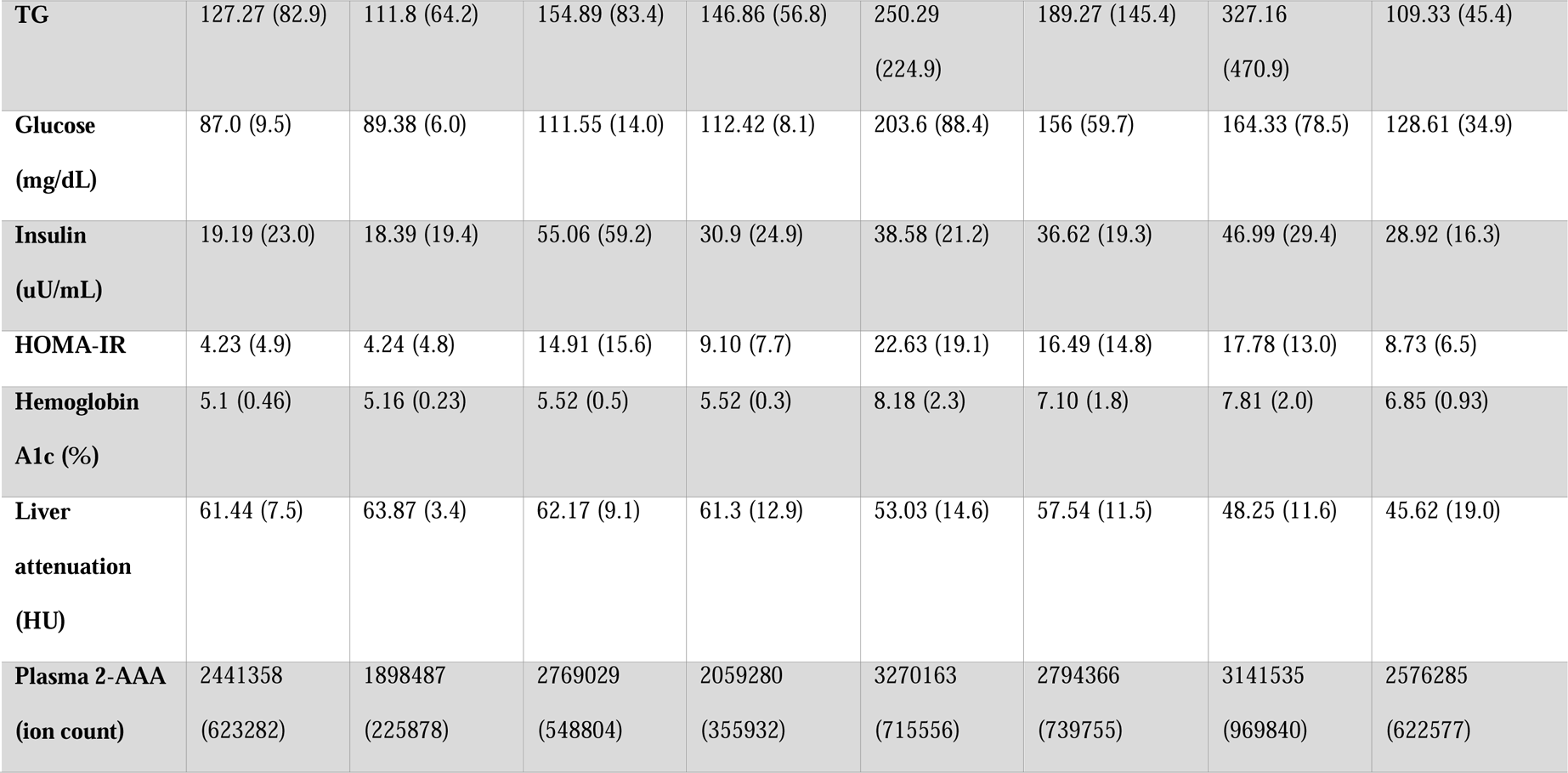
Characteristics of participants of the HATIM Study

### Plasma 2-AAA levels are higher in men than in women, and higher in Asian individuals

There was a significant difference in plasma 2-AAA by sex in the 2-AAA Study, with higher levels in men than in women (plasma 2-AAA 95.99±33.7 vs. 68.43±27.7 ng/ml, P<0.0001; **Figure 1A**). A similar difference by sex was observed in the HATIM Study samples, with higher levels in men than women (plasma 2-AAA ion count 281×10^4^ ± 73 x10^4^ vs. 242 x10^4^ ± 65 x10^4^ ion count, P=0.004; **Figure 1C**). Because other risk factors also differ by sex, we performed stepwise linear regression models including risk factors (BMI, fasting glucose, cholesterol, HDL, LDL, TG), and found that the associations with sex remained significant (P<0.001 *2-AAA Study*, P<0.02 *HATIM Study*). We observed a significant difference by self-reported race in the 2-AAA Study (Overall P=0.002; **Figure 1B**), with individuals self-identifying as Asian having borderline significantly higher plasma 2-AAA (95.68 ± 35.5 ng/ml) compared with individuals self-identifying as Black or African American (72.26 ± 30.0 ng/ml, P=0.05), or white (72.73 ± 30.7 ng/ml, P=0.007). This was not attributable to differences in sex distribution or risk factors between groups. In fact, Asian individuals in the 2-AAA Study had significantly lower BMI (P=0.018) and systolic blood pressure (P=0.005) than other individuals. Interestingly, there was also an overall difference by self-reported race in the HATIM sample (P=0.014; **Figure 1D**), with a trend towards higher levels of 2-AAA in Asian (2-AAA ion count 359 x10^4^±45 x10^4^) compared to Black (2-AAA ion count 249 x10^4^ ± 65 x10^4^) and white (2-AAA ion count 279×10^4^ ± 75×10^4^) individuals, although there were only three individuals self-identifying as Asian in this sample, so the differences did not reach statistical significance in post hoc tests. There was no association between 2-AAA and age in either dataset.

**Figure 1.**
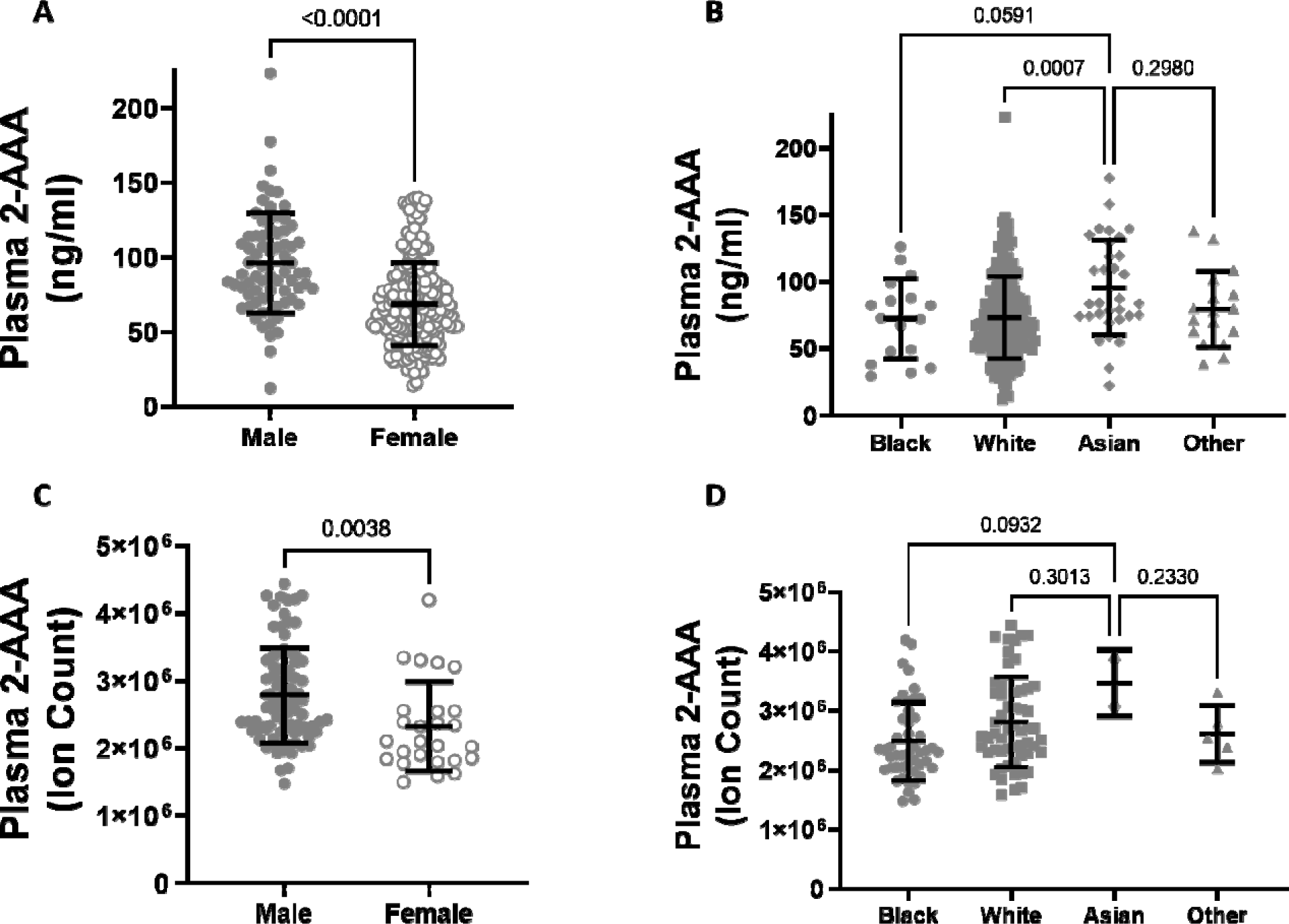
Plasma 2-AAA is significantly higher in men than women in the 2-AAA (A) and HATIM Study (C). 2-AAA is higher in Asian compared to Black or white individuals in the 2-AAA Study (B) with a similar trend in the HATIM Study (D). Data are expressed as ng/ml for data from the 2-AAA Study and ion counts for the HATIM study.

### Plasma 2-AAA levels associate with dyslipidemia in healthy individuals and PWH

Higher plasma 2-AAA was associated with lower HDL cholesterol (2-AAA Study r^2^=0.267, P<0.001; HATIM r^2^=0.579, P<0.001; **Figure 2 A, B**), and higher triglycerides (2-AAA Study r^2^=0.246, P=0.027; HATIM r^2^=0.526, P=0.007; **Figure 2 C, D**). There was no significant association with LDL cholesterol.

**Figure 2.**
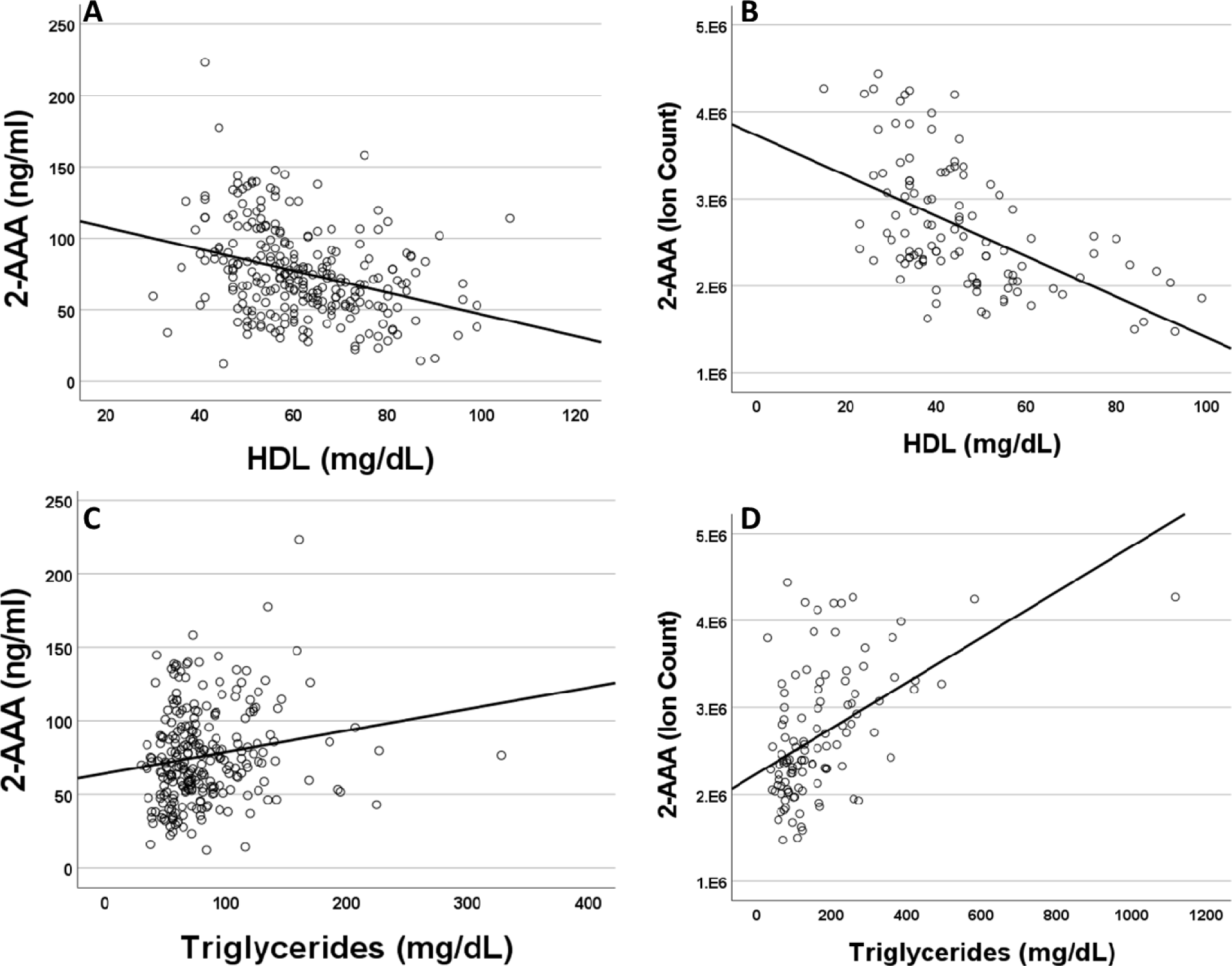
Plasma 2-AAA associates with lower HDL cholesterol and higher Triglycerides in the 2-AAA (A, C) and HATIM (B, D) studies. Data are expressed as ng/ml for data from the 2-AAA Study and ion counts for the HATIM study.

### Higher plasma 2-AAA levels associate with diabetes status in PWH

There were significant differences in plasma 2-AAA by diabetes status within PWH in the HATIM sample (P<0.001, **Figure 3**). Individuals with diabetes had significantly higher levels of 2-AAA (ion count 312×10^4^ ± 75×10^4^) than both the insulin sensitive (ion count 233×10^4^ ± 60×10^4^, P<0.001) and the pre-diabetic (ion count 262×10^4^ ± 58×10^4^, P=0.005) groups in models adjusted for sex, race, BMI and smoking status.

**Figure 3.**
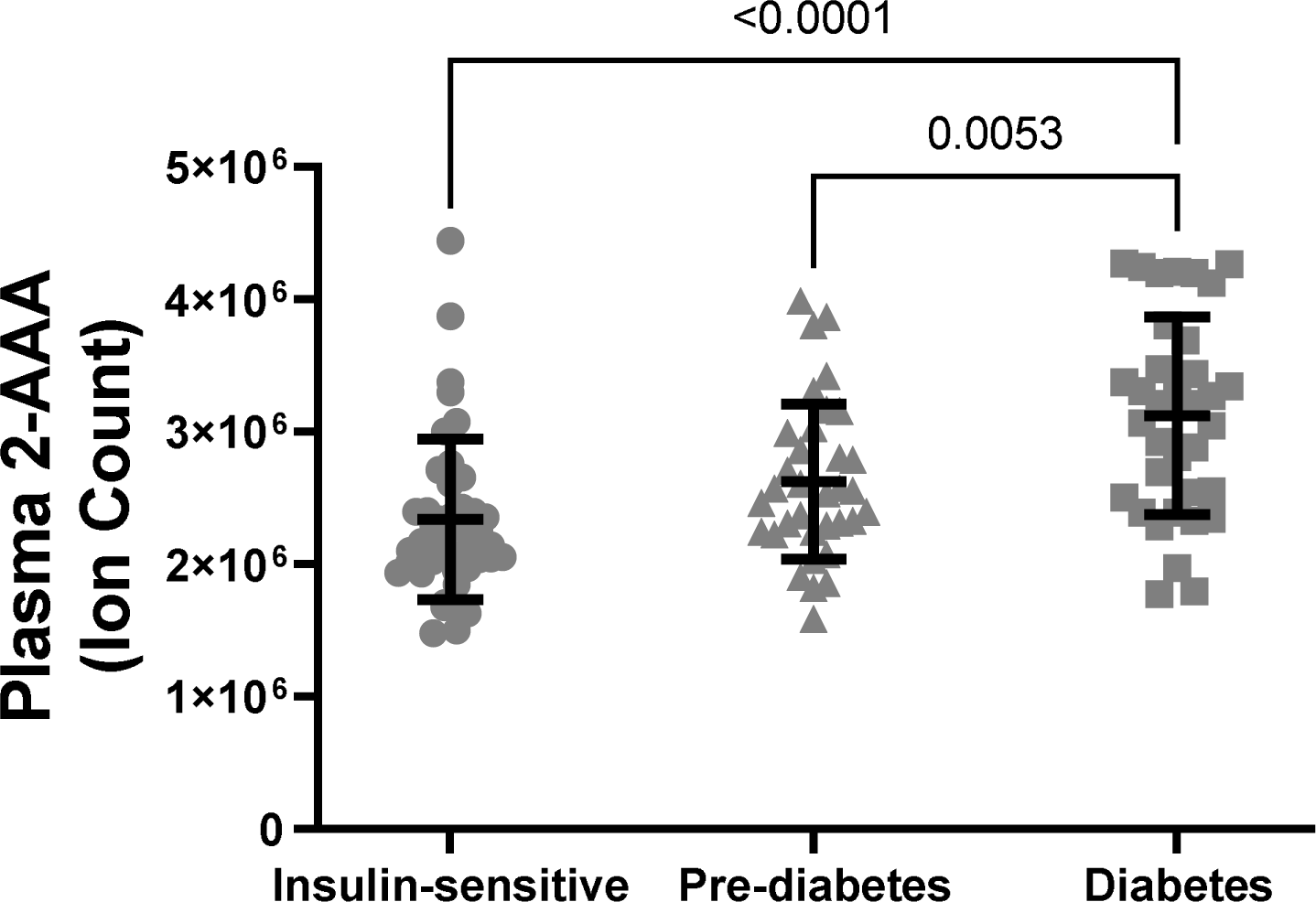
Plasma 2-AAA was significantly higher in PWH and diabetes, compared with PWH who were insulin sensitive or with pre-diabetes.

### Plasma 2-AAA associates with elevated fasting glucose, insulin, and HbA1c in PWH

Across all PWH individuals in HATIM, plasma 2-AAA was associated with increased fasting glucose (r2=0.576, P<0.001), fasting insulin (r2=0.623, P<0.001), HOMA-IR (r2=0.538, P<0.001) and hemoglobin A1c (r2=0.580, P<0.001). In secondary analyses split by diabetes status, 2-AAA associated with glucose and HbA1c only in the individuals with diabetes (P<0.0001 for diabetes, vs P>0.5 for insulin sensitive and pre-diabetes), but 2-AAA was associated with insulin in both people with and without diabetes (P<0.02 insulin sensitive, P<0.002 diabetes). In the 2-AAA Study, a small number of people (n=25) had evidence of potential impaired fasting glucose (IFG, defined as glucose >100mg/dL but <125 mg/dL). While plasma 2-AAA levels were slightly higher within the individuals with IFG (82.5 vs. 75.4 ng/ml), this difference did not reach statistical significance.

### Elevated plasma 2-AAA levels associate with differences in anthropometrics, adipose tissue, and liver density

We found a significant association between plasma 2-AAA and higher BMI in the 2-AAA Study (r^2^=0.275, P<0.001, model adjusted for sex and race), but this was not significant in HATIM. However, in HATIM, higher plasma 2-AAA was significantly associated with increased waist circumference (r^2^=0.219, P<0.001), as well as greater visceral adipose tissue volume (r^2^=0.225, P<0.001), but not with measures of subcutaneous or pericardial adipose tissue. In HATIM, 2-AAA was negatively associated with liver density (r^2^=0.192, P=0.003; **Figure 4**). Lower liver density is a marker of higher proportion of ectopic fat in the liver.

**Figure 4.**
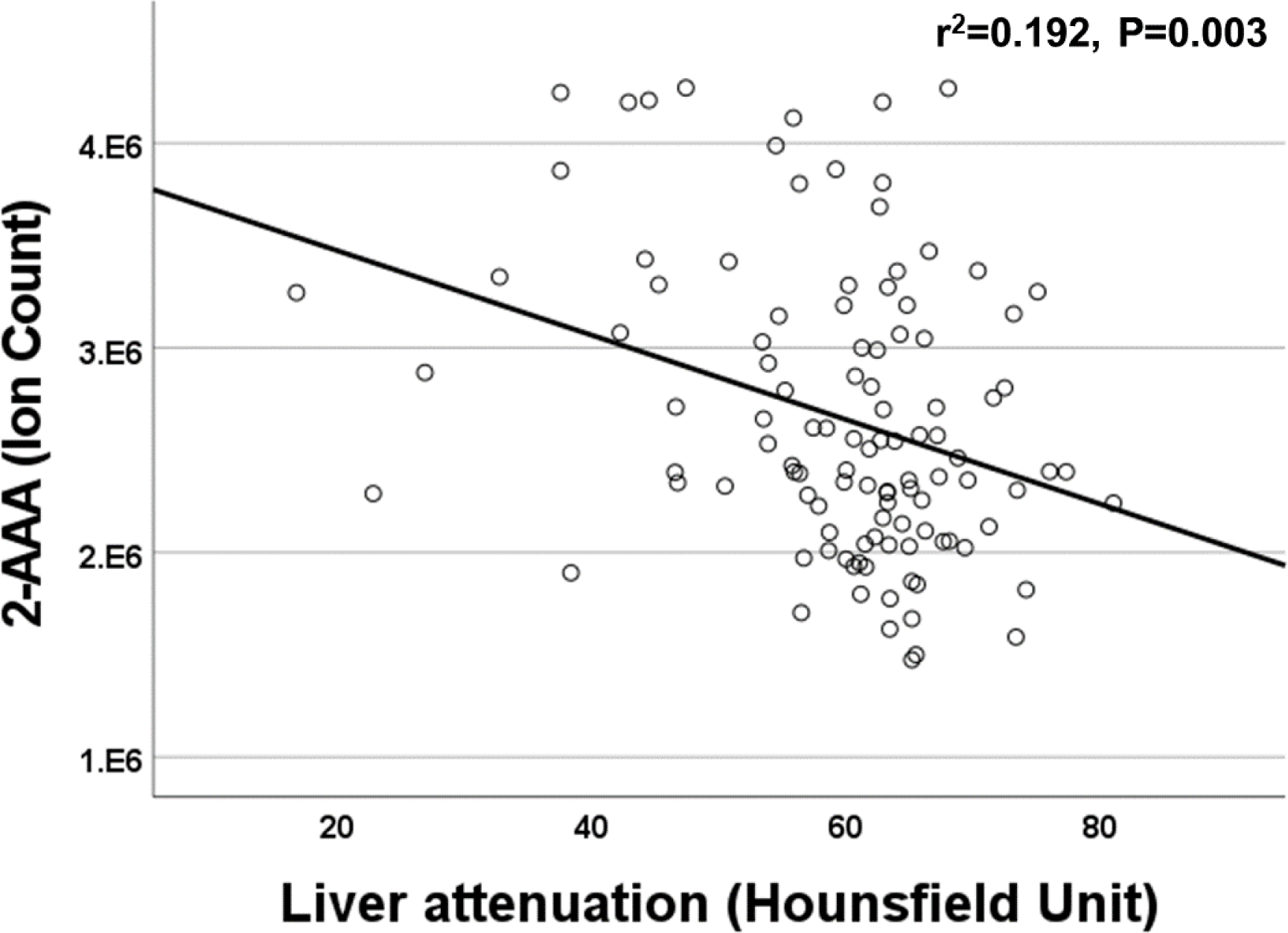
Plasma 2-AAA was negatively associated with liver attenuation in the HATIM Study of PWH.

## DISCUSSION

We measured plasma 2-AAA in two independent samples of individuals across the spectrum of healthy (no diagnosed diseases) to high cardiometabolic risk (diabetes and treated HIV infection). 2-AAA was elevated in diabetes but did not appear to be significantly elevated based on HIV status. We found that plasma 2-AAA is elevated in men compared with women, and in Asian compared with other self-identified ancestries. These associations are constant in both healthy individuals and PWH. We confirmed associations between 2-AAA and both low HDL and high TG, and between 2-AAA and diabetes. We report novel relationships between 2-AAA and visceral adipose tissue measured by CT, and between 2-AAA and higher liver fat. Our data further confirm 2-AAA as an important candidate for further prognostic and therapeutic consideration.

Plasma 2-AAA levels differed by sex, an association that has been reported previously in Mexican young adults (Guevara-Cruz et al., 2018). Men have relatively higher risk of CVD than pre-menopausal women, yet the mechanisms underlying this difference are not fully understood (Tsao et al., 2022). We further report differences by self-reported race, with Asian individuals having higher 2-AAA than other groups. Individuals of Asian ancestry have relatively higher risk of T2D and some CVD given the same risk factor profile as individuals of European ancestry (Ma and Chan, 2013; Buljubasic et al., 2020). The mechanisms underlying this are incompletely understood, and the risk factor profile for CVD in Asians may differ when compared with European ancestry (Paul et al., 2017). While the original discovery of 2-AAA as a diabetes metabolite was in European ancestry (Wang et al., 2013), 2-AAA has also been reported to associate with T2D in Chinese individuals (Wang et al., 2022). Whether differences in 2-AAA may play a role in mediating the relative increased risk in men compared with women, and Asian compared with other ancestries, remains to be determined.

We previously reported that plasma 2-AAA associates with both lower HDL cholesterol and higher triglycerides (Shi et al., 2022). We replicated those associations in the current study, establishing that this relationship is consistent across multiple different samples, including in a cohort of persons with HIV. Based on genetic evidence, 2-AAA drives the decrease in HDL (Shi et al., 2022). While low HDL cholesterol is consistently associated with increased cardiometabolic risk (Castelli et al., 1986; Emerging Risk Factors Collaboration et al., 2009), interventions to alter HDL have shown no benefit (Kingwell et al., 2014). This could be due to differences in HDL composition or function, or due to a causal biomarker that is upstream of HDL. This raises the intriguing hypothesis that elevated 2-AAA, rather than low HDL per se, may be driving increased cardiometabolic risk. However, careful mechanistic studies are required to interrogate this further.

2-AAA was originally discovered as a predictor of diabetes, and is associated with increased insulin secretion in animal models and cells (Wang et al., 2013). In the setting of experimental hyperglycemia in overweight and obese, but otherwise healthy individuals, 2-AAA was significantly decreased following 24 hours of hyperglycemia (Perkins et al., 2019). 2-AAA has been shown to be reduced in the acute setting in response to insulin infusion (Irving et al., 2015). We found that 2-AAA was significantly higher in PWH who have diabetes, than in PWH who were insulin sensitive or pre-diabetic. This is similar to what has been reported in HIV-negative individuals (Wang et al., 2013; Razquin et al., 2019), and suggests that the relationship between 2-AAA and diabetes is consistent across different settings, including against the background of well-controlled HIV infection, a population at increased risk of cardiometabolic disease (Spieler et al., 2022). We found no significant difference in plasma 2-AAA levels based on HIV status in the HATIM cohort within the subset of individuals with diabetes, further suggesting that 2-AAA is a useful biomarker of cardiometabolic risk in multiple at-risk populations. 2-AAA was associated with increased fasting glucose, fasting insulin, and hemoglobin A1c in the HATIM study. However, the association between 2-AAA and glucose was only significant in individuals with diabetes; 2-AAA was not associated with fasting glucose in insulin sensitive individuals in the 2-AAA Study or HATIM, or in individuals with pre-diabetes in HATIM. In contrast, 2-AAA was associated with higher insulin in individuals with and without diabetes. This distinction between the glycemic and insulin axis is consistent with the hypothesis that 2-AAA is an early marker or driver of hyperinsulinemia and is associated with elevated insulin before the development of overt hyperglycemia or diabetes. These data further support a mechanism where elevated 2-AAA precedes the onset of hyperglycemia, and associates with hyperinsulinemia even in individuals who appear insulin sensitive. Associations between 2-AAA and hyperglycemia are likely secondary to insulin resistance. However, further in-depth studies are required to assess potential reciprocal regulation of 2-AAA and insulin.

2-AAA was positively associated with BMI in the 2-AAA study, but not in the HATIM study. However, there was a significant association between 2-AAA and waist circumference in HATIM. This may suggest that the relationship between 2-AAA and adiposity is modulated by HIV-associated effects on adipose distribution (Koethe et al., 2020). Previous studies have also highlighted an association between 2-AAA and obesity, including both BMI and waist circumference (Dugas et al., 2016; Ho et al., 2016; Libert et al., 2018; Lee et al., 2019). While one study has found that 2-AAA is protective against obesity and diabetes in mice (Xu et al., 2019), these findings are in contrast to all other studies, and may be related to specific metabolic anomalies in the mouse model used (Xu et al., 2018; Wang et al., 2021, 1). In our study, 2-AAA associated with increased visceral fat in HATIM, but not subcutaneous or pericardial fat. These data are consistent with a previous study, where 2-AAA was associated with metabolically unhealthy central obesity, compared with metabolically healthy peripheral obesity (Gao et al., 2016). Thus, 2-AAA may relate specifically to pathogenic adipose tissue dysfunction, rather than to obesity itself.

Plasma 2-AAA associated with lower liver density, which corresponds to higher liver fat, and is considered a measure of hepatic steatosis. Previous data in mice found an association between 2-AAA and liver mass (Wu et al., 2014), however, to our knowledge our study describes this for the first time in humans. Elevated 2-AAA may thus be a risk factor for hepatic steatosis and development of fatty liver disease, however, whether this is independent of associations with BMI, visceral fat and circulating lipids remains to be determined.

Our study had several strengths. We analyzed plasma 2-AAA in two separate samples of well-phenotyped individuals, including both healthy individuals and PWH across the diabetes spectrum, allowing us to assess whether the relationship between 2-AAA and cardiometabolic risk markers is consistent in the settings of chronic viral-induced inflammation and in individuals without diagnosed disease.. 2-AAA was not measured in many previous metabolomic studies, and is not consistently detected or reported on popular metabolomics panels (e.g. Metabolon). Thus, the importance of this metabolite in cardiometabolic health may be under-appreciated. We used a targeted assay in the 2-AAA study to quantify 2-AAA, providing important data on circulating levels in healthy individuals. To our knowledge, this is the first study to measure associations between 2-AAA and metabolic disease in PWH. PWH suffer a disproportionate burden of diabetes, hypertension, fatty liver, and dyslipidemia compared to HIV negative persons (Currier et al., 2008; Vodkin et al., 2015; Maurice et al., 2017; Nansseu et al., 2018), and allows for validation of the relevance of 2-AAA to disease within the setting of a highly-inflammatory exaggerated phenotype. Our study also had some limitations. Plasma 2-AAA was measured using a different method in HATIM compared with the 2-AAA study, limiting our ability to directly compare levels of 2-AAA in PWH compared with healthy individuals. However, we were able to compare levels between PWH and HIV-negative within a subset of individuals with diabetes. We also had limited sample size to fully characterize the differences by race across both samples, with small numbers of Black individuals in the 2-AAA study and small numbers of Asian individuals in the HATIM study.

In conclusion, our study establishes differences in plasma 2-AAA by sex and race, confirms associations between 2-AAA and dyslipidemia in both healthy individuals and PWH with or without diabetes, and highlights novel relationships between 2-AAA and liver fat and visceral adipose tissue. Further mechanistic and longitudinal studies are required to establish whether 2-AAA is causally linked to cardiometabolic disease.

## Data Availability

All data produced in the present work are contained in the manuscript

## ACKNOWLEDGEMENTS

We wish to thank the participants of the 2-AAA Study and the HATIM Study. We also thank the work of the Vanderbilt Diet, Body Composition and Human Metabolism core staff.

